# MOCAT: Multi-Omics Integration with Auxiliary Classifiers Enhanced Autoencoder

**DOI:** 10.1101/2023.12.20.23300334

**Authors:** Xiaohui Yao, Xiaohan Jiang, Haoran Luo, Hong Liang, Xiufen Ye, Yanhui Wei, Shan Cong

## Abstract

**Background:** Integrating multi-omics data is emerging as a critical approach in enhancing our understanding of complex diseases. Innovative computational methods capable of managing high-dimensional and heterogeneous datasets are required to unlock the full potential of such rich and diverse data.

**Methods:** We propose a Multi-Omics integration framework with auxiliary Classifiers-enhanced AuToencoders (MOCAT), for comprehensive utilization of both intra- and inter-omics information. Additionally, attention mechanisms with confidence learning are incorporated for enhanced feature representation and trustworthy prediction.

**Results:** Extensive experiments were conducted on four benchmark datasets to evaluate the effectiveness of our proposed model, including BRCA, ROSMAP, LGG, and KIPAN. Our model significantly improved most evaluation measurements and consistently surpassed the state-of-the-art methods. Ablation studies showed that the auxiliary classifiers significantly boosted classification accuracy in both the ROSMAP and LGG datasets. Moreover, the attention mechanisms and confidence evaluation block contributed to improvements in the predictive accuracy and generalizability of our model.

**Conclusions:** The proposed framework exhibits superior performance in disease classification and biomarker discovery, establishing itself as a robust and versatile tool for analyzing multi-layer biological data. This study highlights the significance of elaborated designed deep learning methodologies in dissecting complex disease phenotypes and improving the accuracy of disease predictions.

## 1 Introduction

Recent advancements in omics technologies have enabled large-scale data acquisition across multiple biological layers, including genomics, transcriptomics, proteomics, metabolomics, and many others. Considering that each type of omics data contributes distinct layers of biological information, data integration serves as an efficient tool in multi-omics studies, for not only providing a comprehensive understanding of the multifaceted complexity inherent in biological phenomena but also substantially improves our capabilities in elucidating disease mechanisms and identifying disease biomarkers [1, 2].

Compared to single-omics studies, multi-omics analyses require computational methodologies to encompass a more holistic perspective. These advanced methods aim to overcome the inherent limitations of single-omics approaches by providing nuanced insights into complex biological interactions. However, the complexity and heterogeneity contained within and across multi-omics data pose significant challenges to their integration and downstream tasks. Traditional approaches for analyzing multi-omics data mainly rely on statistical methodologies, as indicated in studies [3–5]. These statistical approaches often encounter difficulties in feature extraction, typically requiring manual intervention that can be both labor-intensive and suboptimal for interpreting global feature significance. Deep learning models have been widely designed to address this issue and demonstrated superior predictive capabilities and proficiency in identifying nonlinear and hierarchical features [6–8].

Deep neural networks in multi-omics analyses enable the autonomous extraction of relevant features and facilitate the identification of intricate associations among them. However, when applying neural networks to multi-omics data, a series of challenges remain, one notable issue being the ‘curse of dimensionality’ [9]. Due to the multiple causes and pathogenic mechanisms underlying complex diseases, omics data are particularly prone to this issue. Such high dimensionality creates intricate spatial distributions that can impede both traditional machine learning and deep learning algorithms in their classification tasks. Preserving all original features magnifies the computational complexity in such high-dimensional spaces, inducing overfitting and diminishing the predictive accuracy. To address these challenges, autoencoder models have been investigated as a means to transform and integrate multi-omics features, particularly for discerning disease subtypes [10–12]. Various strategies based on autoencoders have been proposed to integrate high-dimensional, multi-source datasets and to derive low-dimensional latent representations. For example, Wang et al. [13] employed autoencoders to align and integrate data from single-cell RNA-seq and ATAC-seq, adeptly mapping the sparse and noisy data from varied spaces into a harmonized subspace for improved alignment and integration. Lin et al. [14] introduced scMDC, an architecture featuring one encoder for cascading data and two decoders for each data modality. This design uniquely characterizes distinct data sources and co-learns deep embeddings of latent features for cluster analyses. Autoencoders use a combination of nonlinear functions to reconstruct the original inputs, which can be used as new feature representations of the original data. These algorithms have been proven effective in producing clinically relevant features [15], analyzing high-dimensional omics data [16, 17], and integrating multi-omics data [7, 18]. However, autoencoders can exhibit suboptimal performance in certain tasks, especially when the generated subrepresentations are utilized for downstream tasks [7, 19]. This issue often originates from the focus of traditional autoencoder objective functions on input reconstruction, which can limit their effectiveness in classification tasks.

In addition to omics-specific feature extraction, data fusion is another key step in multi-omics studies. Attention mechanisms have been a robust technique for enhancing classification performance by selectively emphasizing salient features across various omics levels. These mechanisms enhance the model performance by prioritizing more relevant features for classification outcomes. Such adaptability is particularly crucial in light of the varying importance of different features. As substantiated by feature interpretation studies such as [20], attention mechanisms are justified in their application for multi-omics fusion, given their capacity to weigh the importance of features in diagnosis prediction adaptively.

On the other hand, deep learning models for classification tasks typically adopt maximum class probability (MCP), that is, the highest probability values given by the softmax output, to evaluate the prediction confidences [21]. This can lead to assigning high confidence values to even incorrect predictions. To mitigate this limitation and enhance classification accuracy, the true class probability (TCP) criterion is integrated into the loss function [22, 23]. Unlike MCP, TCP assesses the predicted probability for each class against the probability of the true class label, incorporating these values into the loss computation. This criterion acts as a regularizer during training by offering more detailed insights into the performance of the classifier on individual sample predictions. This becomes more essential in challenging scenarios, like those where performance improvement is hindered due to hard samples (that is commonly observed in complex diseases) or when only a few samples are incorrectly predicted due to the well-designed models.

Upon recognizing the observed limitations in existing methods, we present a Multi-Omics integration framework with auxiliary Classifiers-enhanced AuToencoder (MOCAT) to improve both stability and predictive accuracy in disease classification tasks. Acknowledging the importance of explicability within the domain of biomedical research, our framework also incorporates model interpretability for biomarker discovery. The architecture employs autoencoders for efficient high-dimensional feature compression, while the integration of omics-specific classifiers promotes refined optimization aligned with disease prediction. Furthermore, adopting attention mechanisms affords greater flexibility in fusing multiple omics types. We also incorporate the trustworthy strategy to facilitate fine-grained optimization in the weighting of the classification network, culminating in an augmented accuracy of classification. Benchmark experiments and comparative evaluations show that the proposed model outperforms existing state-of-the-art methods, with extensive validations demonstrating both the reliability and interpretability of the proposed framework.

Our main contributions are summarized as the following:

- **Omics-Specific Feature Optimization:** We introduce auxiliary classifiers tailored for each type of omics data, which significantly enhances feature representation by identifying the most informative biomarkers pertinent to disease states.
- **Enhanced Classifier Confidence Calibration:** We incorporate the true class probability criterion to regularize classifier confidence of incorrect predictions, thereby improving model overconfidence and enhancing predictive accuracy.
- **Explainability:** By integrating mechanisms that elucidate the decision-making process of our model, we provide predictive proficiency and facilitate a deeper understanding of the underlying biological phenomena, thereby aiding in the interpretive aspects of biomarker discovery.
- **State-Of-The-Art (SOTA) Performance:** The proposed framework has yielded superior results on four independent datasets, indicating an improvement over the current benchmarks.

## 2 Materials and Methods

### 2.1 Datasets

To conduct fair comparative experiments, we adopt public data preprocessed by Wang et al. [20], which provide four datasets for multi-omics disease classification, including a binary classification ROSMAP dataset for Alzheimer’s disease (AD) patients and normal controls (NC), a BRCA dataset for PAM50 subtype classification of invasive breast cancer (five-class), an LGG dataset for the grade classification of gliomas (binary), and a KIPAN dataset for subtype classification of renal cancer (three-class). Table 1 shows the detailed information of these datasets, where the preprocessed features were used for training.

**Table 1.** Summary of datasets.

| Dataset | Sample | Number of raw features |  |  | Number of features for training |  |  |
| --- | --- | --- | --- | --- | --- | --- | --- |
|  |  | mRNA | methy | miRNA | mRNA | methy | miRNA |
| ROSMAP | NC: 169, AD: 182 | 55,889 | 23,788 | 309 | 200 | 200 | 200 |
| BRCA | Luminal A: 436, Luminal B: 147,<br>HER2-enriched: 46, Normal-like: 115, Basal-like: 131 | 20,531 | 20,106 | 503 | 1,000 | 1,000 | 503 |
| LGG | Grade 2: 246, Grade 3: 264 | 20,531 | 20,114 | 548 | 2,000 | 2,000 | 548 |
| KIPAN | KICH: 66, KIRC: 318, KIRP: 274 | 20,531 | 20,111 | 445 | 2,000 | 2,000 | 445 |

### 2.2 Model Formulation

The proposed model is structured into three sequential phases for comprehensive data analysis. Phase 1 focuses on efficiently compressing high-dimensional data to extract critical omics-specific features. Phase 2 involves the integration of multi-omics data and confidence-based disease prediction. Finally, Phase 3 is concerned with biomarker discovery, harnessing the insights collected from the previous phases. The overall architecture of the proposed model is shown in Figure. 1.

**Fig. 1.**
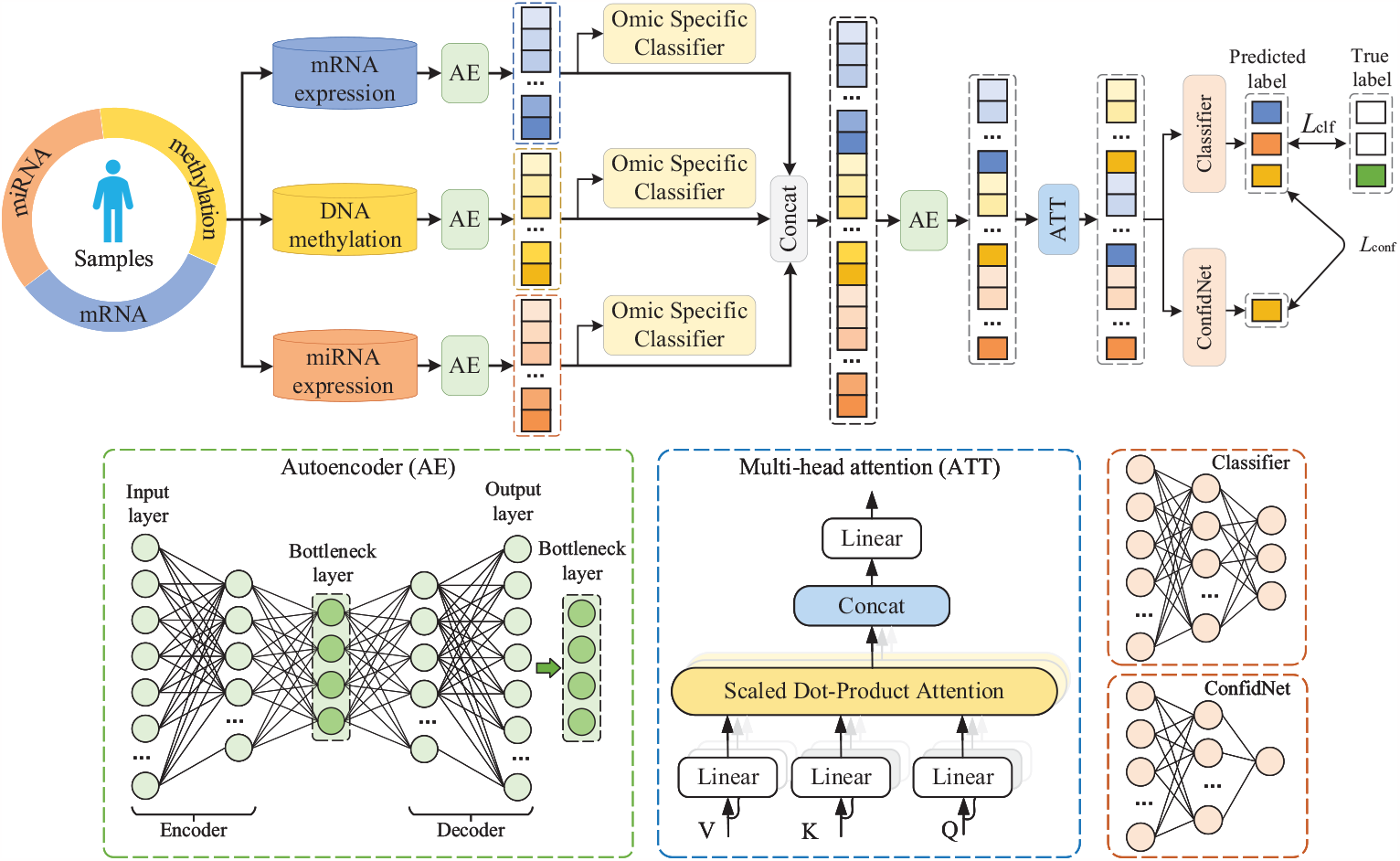
Framework of the MOCAT. The top panel shows the overall architecture of the proposed model: 1) high-dimensional features of multiple omics datasets are fed into an autoencoder network for dimensionality reduction to obtain representative features; 2) three omics-specific auxiliary classifiers are trained to assist in learning more compact and accurate sub-representations; 3) feature sub-representations are fused and further compressed by an autoencoder network a self-attention module to fuse the complementary information embedded across multi-omics adaptively; 4) confident network (ConfNet) is employed to adjust the prediction confidence linked to the fused features adaptively. The bottom panel illustrates the detailed architectures of the autoencoder, attention, and confidence networks.

#### 2.2.1 Phase 1: Omics-Specific Feature Extraction

In phase 1, high-dimensional features of each omics dataset are fed into autoencoder networks for extracting representative features. At the same time, each omics data is separately trained to assist the autoencoder network in learning a more compact and accurate representation. Developing independent models for each omics dataset can help avoid losing the specificity of each data source, as they may exhibit distinct dynamics. The independent models are expected to provide reliable feature information for multimodal fusion.

##### Autoencoders for dimensionality reduction

Three autoencoders are trained separately on different omics types. In particular, each autoencoder uses a multi-objective optimization method in the encoder with Dropout, BatchNorm, and ELU activation functions to compress the original data and extract corresponding low-dimensional representations. The Dropout layer is generally set after the fully connected layer and randomly drops part of the nodes according to a preset ratio during training, effectively improving the problem of model overfitting and improving model generalization. The BatchNorm layer, as shown in Eq. 1, is used to normalize the mean and variance of features and is widely applied in deep learning tasks. This approach speeds up model training and improves model stability while alleviating issues like gradient vanishing and gradient explosion.

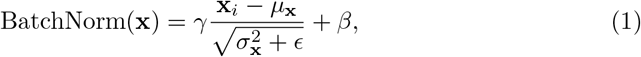

where *μ*_**x**_ and 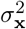 are the batch mean and variance, *γ* is a scale parameter used to scale the normalized data, *ϵ* is a small constant added to the denominator to prevent division by zero, and *β* is a shifting parameter used to shift the normalized results by the batch mean.

The ELU activation function can better manage the gradient vanishing problem, making the training converge faster, thus achieving better results:

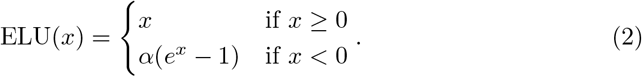

In detail, each autoencoder model consists of multiple fully connected layers, with a bottleneck layer in the middle that minimizes node size. The intermediate layer with the fewest nodes serves as the bottleneck layer for dimensionality reduction in the original dataset. The last layer reconstructs the raw data from the first layer. We aim to minimize reconstruction errors and extract better feature representations at the bottleneck layer. The effectiveness of feature representation can be evaluated by omics-specific classifiers.

##### Omics-specific classifiers

Auxiliary classifiers specific to each omics data are incorporated to fulfill two objectives: (i) improve representation by encouraging the autoencoders to learn more fine-grained and discriminative features; (ii) improve prediction performance by forcing the model to learn from multiple perspectives. The classification loss from the omics-specific classifiers directs the autoencoders in refining feature compression, ensuring that the compressed representations align with the distinguishing characteristics of each omics data. This alignment is intuitively thought to promote the overall effectiveness of the autoencoder for dimensionality reduction.

Overall, *M* omics-specific feature subrepresentations F^(*m*)^, *m* ∈ {1, …, *M*} were obtained from phase 1. The loss of the first phase includes the reconstruction loss 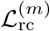 of each autoencoder and the auxiliary classification loss 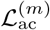 of each omics-specific classifier, and can be expressed as:

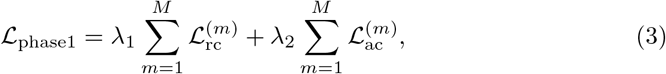

where *λ*_1_ and *λ*_2_ are hyperparameters for adjusting different losses. We set *λ*_1_ = 1 and *λ*_2_ = 0.005 in our experiment.

#### 2.2.2 Phase 2: Cross-Omics Fusion and Trustworthy Prediction

Integrating heterogeneous multi-omics data, characterized by varying expression patterns and dimensions, presents a significant challenge. In phase 2 of our approach, we delve into improving both multi-omics fusion and final disease prediction, utilizing feature representations derived from phase 1. We specifically apply the attention mechanism to establish global correlations within the fused features and introduce the classification confidence mechanism into the network to enhance its prediction performance.

##### Adaptive fusion with autoencoder and attention

We first concatenated the omics-specific subrepresentations obtained from phase 1 to create the preliminary fused representations. Subsequently, an autoencoder network was trained to map these heterogeneous features into a novel embedding space. This space is designed to learn and encapsulate shared representations across the different omics data types. This procedure facilitates the extraction of discriminative and representative features from a more comprehensive perspective, thereby augmenting the overall effectiveness of the model. The output **Z**_AE_ of the autoencoder layer is calculated as follows:

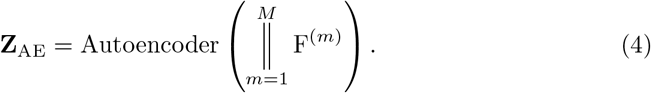

The integrated features were subsequently fed into an attention layer. This is designed to capture and emphasize the distinct significance of various omics modalities, thereby augmenting the efficacy of our model. It has been noted that different types of omics data contribute variably to the aggregate predictive accuracy. The integration of the attention mechanism allows for a dynamic recalibration of the influence exerted by the fused modality features during the classification procedure. Given the input **Z**_AE_, the output **Z**_Att_ of the attention layer is calculated as follows:

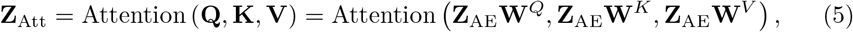

where 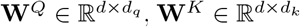, and 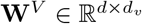 with *d*_*q*_ = *d*_*k*_ are three learnable weight matrices for generating the corresponding matrices of query 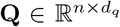, key 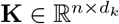, and value 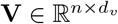, *n* is the number of samples and *d* is the embedding dimensionality of the previous autoencoder layer.

In summary, autoencoders and self-attention can promote and assist each other in optimization. This collaboration is particularly important in addressing the short-comings of conventional autoencoders, which may not sufficiently retain cross-omics information. The combination of autoencoder and attention networks can promote the adaptive fusion of multi-omics features, therefore maximizing the utilization of complementary information embedded across multi-omics data.

##### Trustworthy prediction with ConfNet

In addition to improving the representation effectiveness of intra- and inter-omics data, we also employed the trustworthy strategy to assess and adaptively adjust the prediction confidence linked to the fused features.

The traditional method for determining confidence in classification, known as the maximum class probability (MCP), relies on the highest probability output of the softmax function. For a given input feature matrix **Z**_Att_, the classifier acts as a probabilistic model. This model assigns a predictive probability distribution *P* (*Y* |**Z**_Att_) for each class in the set *k* = {1, …, *K*}. The class with the highest probability is then selected as the predicted class, denoted as *ŷ*= arg max_*k*∈{1,…,*K*}_ *P* (*Y* = *k* |**Z**_Att_). However, a notable issue with MCP is its tendency to exhibit overconfidence for incorrect predictions.

The true class probability (TCP) confidence criterion was proposed to solve this problem [22], by assigned confidences according to *P* (*Y* = **y**^*^ |**Z**_Att_), where **y**^*^ represents the true label vector. TCP and MCP yield equivalent results when a sample is correctly classified. However, for misclassified samples, TCP provides a more conservative and, thus, potentially more accurate confidence value. The direct estimation of TCP confidence on the test set is not feasible due to the absence of true labels. To solve this problem, a confidence network (denoted as ConfidNet) was introduced to the training data, and the parameters were learned as follows:

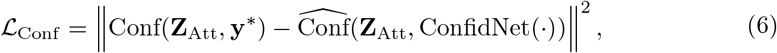

where Conf(**Z**_Att_, **y**^*^) is the TCP confidence we propose to learn. As illustrated in Figure. 1, both the confidence network and the classifier were built upon the output of the attention layer. The classifier was trained using the cross-entropy loss and fixed, after which the confidence network was trained according to Eq. 6. In this way, the model is designed to adjust the feature weights in response to misclassified samples adaptively. This dynamic penalization mechanism enhances the model to learn from errors and refine its predictive accuracy. Furthermore, by effectively distinguishing false predictions from true ones through enhanced confidence separation, the TCP criterion holds the potential to boost the generalizability of the model and thereby reduce the risk of overfitting.

Therefore, the loss of phase 2 consists of the reconstruction loss of the fused features ℒ_rc_ and the confidence loss ℒ_conf_ :

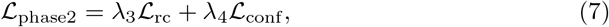

where *λ*_3_ and *λ*_4_ are hyperparameters used to balance different losses and are set to 0.5 in the experiments.

In total, the loss of the entire model includes the phase 1 loss, phase 2 loss, and the cross-entropy loss for the final classification ℒ_clf_:

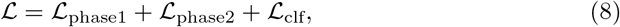

#### 2.2.3 Phase 3: Biomarkers Identification

Identifying biomarkers is fundamental for understanding underlying biological mechanisms and interpreting outcomes in biomedical contexts. Discovering biomarkers via deep learning models facilitates the discernment of highly representative and predictive features in classification tasks. We evaluated the importance of each omics feature to find those showing significant effects on the prediction performance of the model. Specifically, feature ablation was employed wherein each feature was eliminated, and the feature-level importance score was calculated according to the decreasing accuracy. In practice, we repeated five times to obtain the mean importance measurements to reduce experimental variability.

## 3 Results

We evaluated the performance of our proposed method by comparing it with state-of-the-art multi-omics classification approaches using four public datasets. Furthermore, extensive ablation studies were executed to elucidate the efficacy of each component within our framework. We focused on three metrics for binary classification: classification accuracy (ACC), F1 score, and area under the ROC curve (AUC). For multiclass classification datasets, we also focused on three metrics including accuracy (ACC), weighted average F1 score (F1_w), and macroaverage F1 score (F1_m).

### 3.1 Diseases Prediction Comparison

Our comparative analysis encompassed fourteen computational methods, including six early-stage single-omics benchmark algorithms, namely, K-nearest neighbors (KNN) [24], support vector machine (SVM) [25], Lasso [26], random forest (RF) [27], eXtreme Gradient Boosting (XGboost) [28], and fully connected neural networks (NN) [29]. We also evaluated seven advanced multi-omics classification frameworks, which include group-regularized ridge regression (GRridge) [30], Bayesian partial least squares discriminant analysis-based BPLSDA [31], BSPLSDA [31], Concatenate Fusion (CF) for post-modality connection of multi-omics representations [32], Gate Modulated Unit (GMU) for information fusion with gating mechanisms [33], and the two state-of-the-art algorithms Mogonet [20] and Dynamic [34]. For fair comparisons, we evaluated all methods according to the same experimental settings as Mogonet [20], and the outcomes were expressed as the mean and standard deviation of five experiments.

As shown in Table 2 and Table 3, our model outperformed both the benchmark and state-of-the-art methods on both binary and multiclass classification tasks. Our approach consistently outperformed existing methods on the ROSMAP, BRCA, and LGG datasets, demonstrating the robustness and adaptability of our model. In the case of the KIPAN dataset, performance from our model was on par with advanced algorithms, validating its competitive capability. Statistical analysis demonstrated that the performance improvements are significant (*P <* 0.05), further confirming the substantial superiority of the proposed model.

**Table 2.** Comparison with state-of-the-art methods on ROSMAP and BRCA datasets.

| Method | ROSMAP (2 Categories) |  |  | BRCA (5 Categories) |  |  |
| --- | --- | --- | --- | --- | --- | --- |
|  | ACC(%) | F1(%) | AUC(%) | ACC(%) | F1_w(%) | F1_m(%) |
| KNN | 65.7±3.6 | 67.1±4.4 | 70.9±4.5 | 74.2±2.4 | 73.0±2.3 | 68.2±2.5 |
| SVM | 77.0±2.4 | 77.8±1.6 | 77.0±2.6 | 72.9±1.8 | 70.2±1.5 | 64.0±1.7 |
| Lasso | 69.4±3.7 | 73.0±3.3 | 77.0±3.5 | 73.2±1.2 | 69.8±1.5 | 64.2±2.6 |
| RF | 72.6±2.9 | 73.4±2.1 | 81.1±1.9 | 75.4±0.9 | 73.3±1.0 | 64.9±1.3 |
| XGBoost | 76.0±4.6 | 77.2±4.5 | 83.7±3.0 | 78.1±0.8 | 76.4±1.0 | 70.1±1.7 |
| NN | 75.5±2.1 | 76.4±2.1 | 82.7±2.5 | 75.4±2.8 | 74.0±3.4 | 66.8±4.7 |
| GRridge | 76.0±3.4 | 76.9±2.9 | 84.1±2.3 | 74.5±1.6 | 72.6±1.9 | 65.6±2.5 |
| BPLSDA | 74.2±2.4 | 75.5±2.3 | 83.0±2.5 | 64.2±0.9 | 53.4±1.4 | 36.9±1.7 |
| BSPLSDA | 75.3±3.3 | 76.4±3.5 | 83.8±2.1 | 63.9±0.8 | 52.2±1.6 | 35.1±2.2 |
| CF | 78.4±1.1 | 78.8±0.5 | 88.0±0.5 | 81.5±0.8 | 81.5±0.9 | 77.1±0.9 |
| GMU | 77.6±2.5 | 78.4±1.6 | 86.9±1.6 | 80.0±3.9 | 79.8±5.8 | 74.6±5.8 |
| Mogonet | 81.5±2.3 | 82.1±2.2 | 87.4±1.2 | 82.9±1.8 | 82.5±1.6 | 77.4±1.7 |
| Dynamic | 84.2±1.3 | 84.6±0.7 | 91.2±0.7 | 87.7±0.3 | 88.0±0.5 | 84.5±0.5 |
| MOCAT (Ours) | **87.6±0.7\*** | **87.5±0.6\*** | **92.3±0.9\*** | **88.5±0.3\*** | **88.9±0.3\*** | **86.2±0.7\*** |

Note: Mean and standard deviation are presented, and the best results are in bold. Compared to the suboptimal model, the superior model is denoted by \* to indicate a statistically significant improvement ( $P < 0.05$ ) when using the two-sample t-test.

**Table 3.**
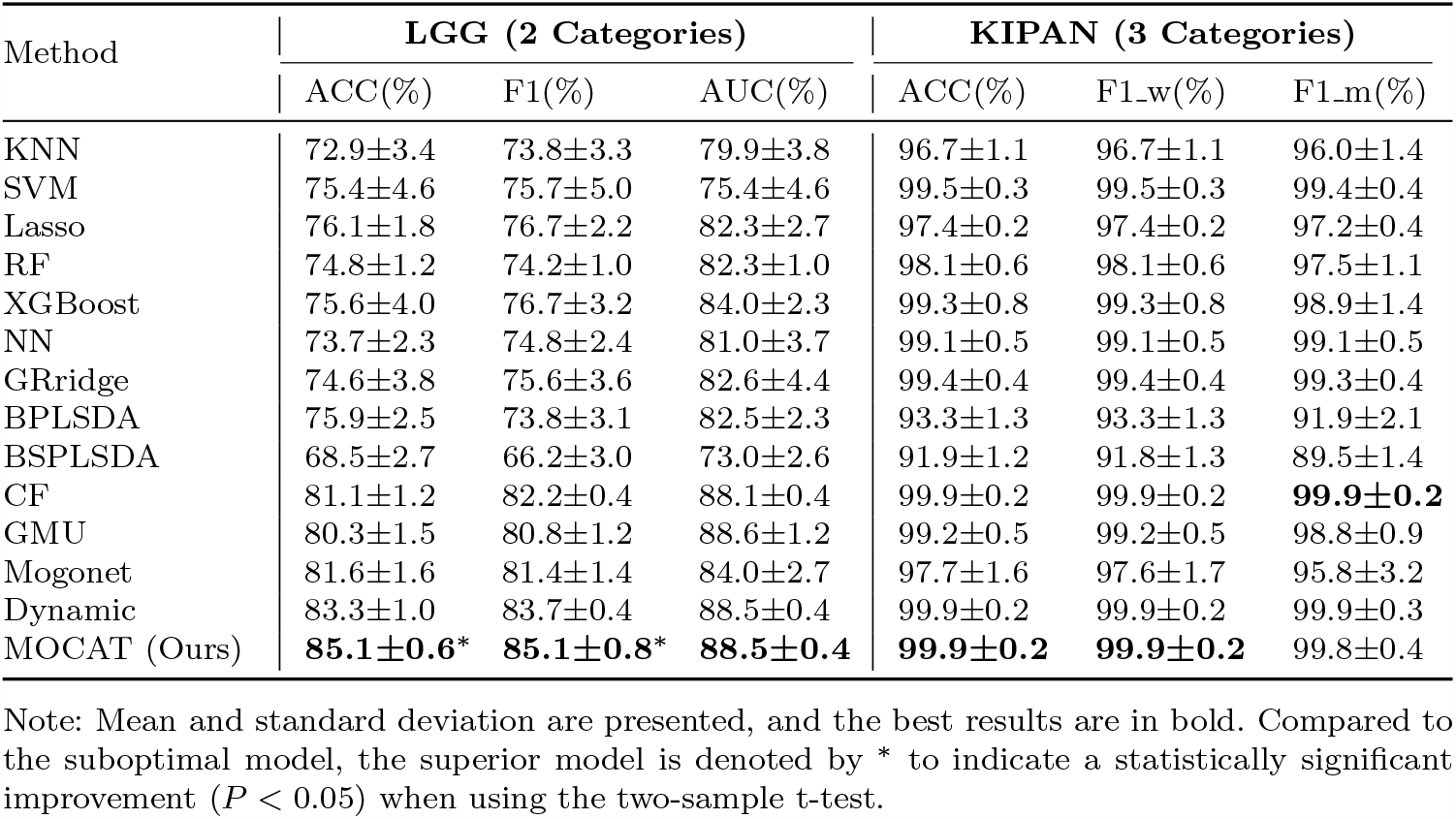
Comparison with state-of-the-art methods on LGG and KIPAN datasets.

### 3.2 Comparative Analysis Across Varied Omics Types

In our investigation, we endeavored to integrate diverse omics data types—specifically mRNA, DNA methylation, and miRNA—to provide a more comprehensive understanding of disease etiology and to enhance the accuracy of disease classification beyond what is possible with single or dual omics data sources. This is based on the hypothesis that each data contributes uniquely to the model and that integrating multiple sources can lead to more robust performance. We designed a series of ablative studies on the ROSMAP, BRCA, and LGG datasets to validate our hypothesis, excluding the KIPAN dataset due to its relatively straightforward classification nature. We assessed the performance impact when transitioning from using individual omics datasets to combinations of two and ultimately incorporating all three.

Figure. 2 illustrates the performance comparison of using various omics combinations. It can be observed that utilizing all three omics types yielded the highest performance across all three tasks, except the AUC of mRNA+miRNA on the LGG dataset (89.9% > 88.5%). This emphasizes the advantage of harnessing multiple omics, which provide a more comprehensive spectrum of crucial information. Furthermore, it validates the capacity of our proposed model in effectively extracting and integrating representative features from these diverse omics sources.

**Fig. 2.**
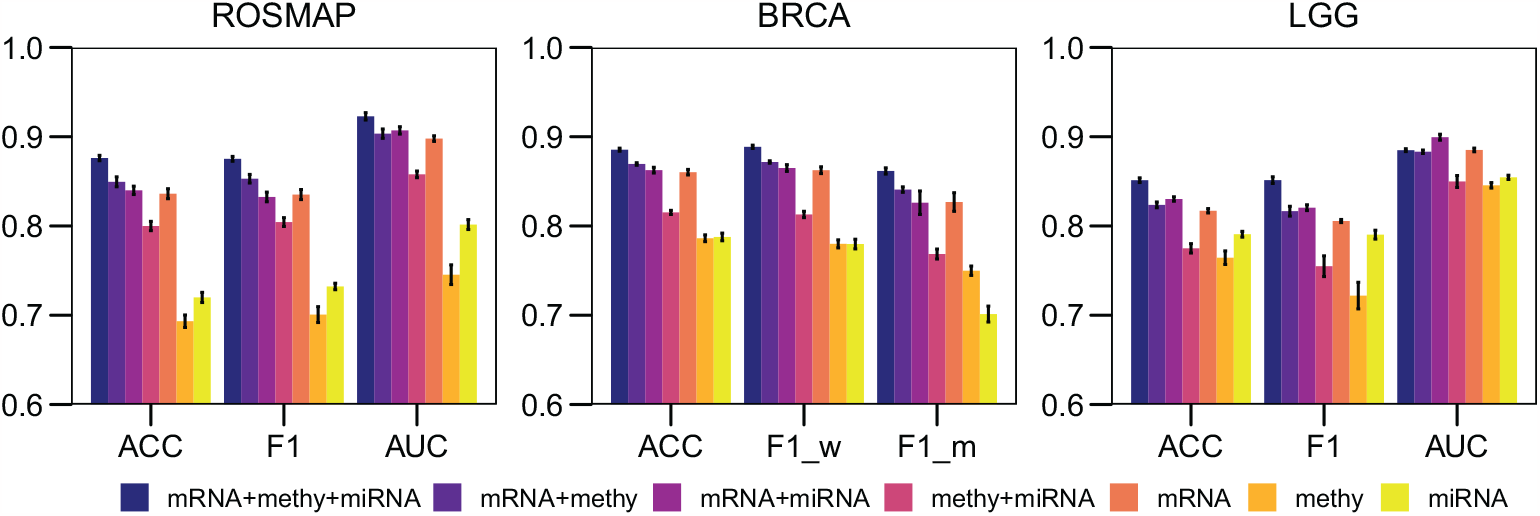
Performance comparisons of different omics combinations using the proposed method. Means and standard errors are presented.

### 3.3 Ablation Study

We performed ablation studies to assess the key modules used in our method, including the omics-specific auxiliary classifiers (AC), the attention mechanism (Att), and the trustworthy strategy (ConfNet). We respectively removed these three components from the proposed model and explored the prediction performance.

Results are summarized in Table 4. We can observe that each critical component contributes to enhancing the classification efficacy of our model. Specifically, the removal of the expressly designed auxiliary classifiers results in a significant decline in performance. This is particularly noticeable in the context of binary classification tasks. Notably, in the ROSMAP and LGG datasets, we obtain an improvement of 8.2% and 5.1% in accuracy and 7.9% and 6.9% in the F1 score, respectively. This underscores the efficacy of omics-specific classifiers in enriching the capacity of autoencoders for nuanced feature representation.

**Table 4.** Ablation study of the key modules. Mean and standard deviation are presented, and the best results are in bold. The overline denotes the ablation of the corresponding module. The asterisk ^*^ denotes a statistically significant difference between the scenarios with and without the respective key module, as computed by the two-sample t-test (*P* < 0.05).

| Dataset | Method | ACC (%) | F1 (%) | AUC (%) |
| --- | --- | --- | --- | --- |
| ROSMAP | $\overline{AC}$: AE<sub>os</sub>+AE<sub>f</sub>+Att+ConfNet | 79.4±2* | 79.6±2.4* | 88.2±0.7* |
| | $\overline{Att}$: AE<sub>os</sub>+AC+AE<sub>f</sub>+ConfNet | 86.7±0.7 | 86.5±0.6 | **92.3±0.4** |
| | $\overline{ConfNet}$: AE<sub>os</sub>+AC+AE<sub>f</sub>+Att | 85.5±0.8* | 85.6±0.8* | 92.2±0.7 |
|  | Ours | **87.6±0.7** | **87.5±0.6** | 92.3±0.9 |
| LGG | $\overline{AC}$: AE<sub>os</sub>+AE<sub>f</sub>+Att+ConfNet | 80.0±1.2* | 78.2±1.4* | 88.8±0.6 |
| | $\overline{Att}$: AE<sub>os</sub>+AC+AE<sub>f</sub>+ConfNet | 83.6±0.5* | 83.4±0.7* | 88.9±0.5 |
| | $\overline{ConfNet}$: AE<sub>os</sub>+AC+AE<sub>f</sub>+Att | 84.1±0.5* | 83.8±0.8* | **89.0±0.4** |
|  | Ours | **85.1±0.6** | **85.1±0.8** | 88.5±0.4 |
| Dataset | Method | ACC (%) | F1<sub>w</sub> (%) | F1<sub>m</sub> (%) |
| BRCA | $\overline{AC}$: AE<sub>os</sub>+AE<sub>f</sub>+Att+ConfNet | 87.8±0.5 | 88.0±0.7 | 84.5±1.7 |
| | $\overline{Att}$: AE<sub>os</sub>+AC+AE<sub>f</sub>+ConfNet | 87.4±0.5* | 87.7±0.5* | 84.8±0.1* |
| | $\overline{ConfNet}$: AE<sub>os</sub>+AC+AE<sub>f</sub>+Att | 87.9±0.3* | 88.1±0.3* | 85.0±0.5* |
|  | Ours | **88.5±0.4** | **88.9±0.4** | **86.2±0.8** |

**Abbreviations.** AC: auxiliary classifiers; AE<sub>os</sub>: omics-specific autoencoders; AE<sub>f</sub>: autoencoders applied on the fused features; Att: self-attention; ConfNet: confidence network.

Incorporating the attention mechanism also enhances almost all of the three experiments. For example, the attention module significantly improves the classification of BRCA subtypes across all three evaluation metrics. This highlights the capacity of the attention mechanism to fine-tune the ability of the model to discern and prioritize critical features across the various omics datasets.

The novel confidence criterion also illustrates increased prediction performance compared to the conventional MCP strategy. The results consistently show that integrating the TCP criterion contributes to performance enhancements in most experiments. Specifically, on the BRCA dataset, the application of TCP results in significant improvements in ACC, AUC, and F1 scores (t-test *P <* 0.05). The ROSMAP and LGG datasets, including the confidence networks, also achieve significantly higher accuracy and F1 scores.

We further monitored the progression of training and testing losses over increasing epochs to investigate if the TCP-based confidence network can help reduce the risk of overfitting. Figure. 3 illustrates the learning curve comparison of the ROSMAP classification task. It shows that the model without using ConfNet exhibits tendencies toward overfitting (around epoch 300), while a limitation is effectively reduced by including the TCP criterion. This suggests that the novel confidence criterion can improve prediction performance and contribute to the generalization capabilities of the model, making it more robust and reliable when applied to unseen data.

**Fig. 3.**
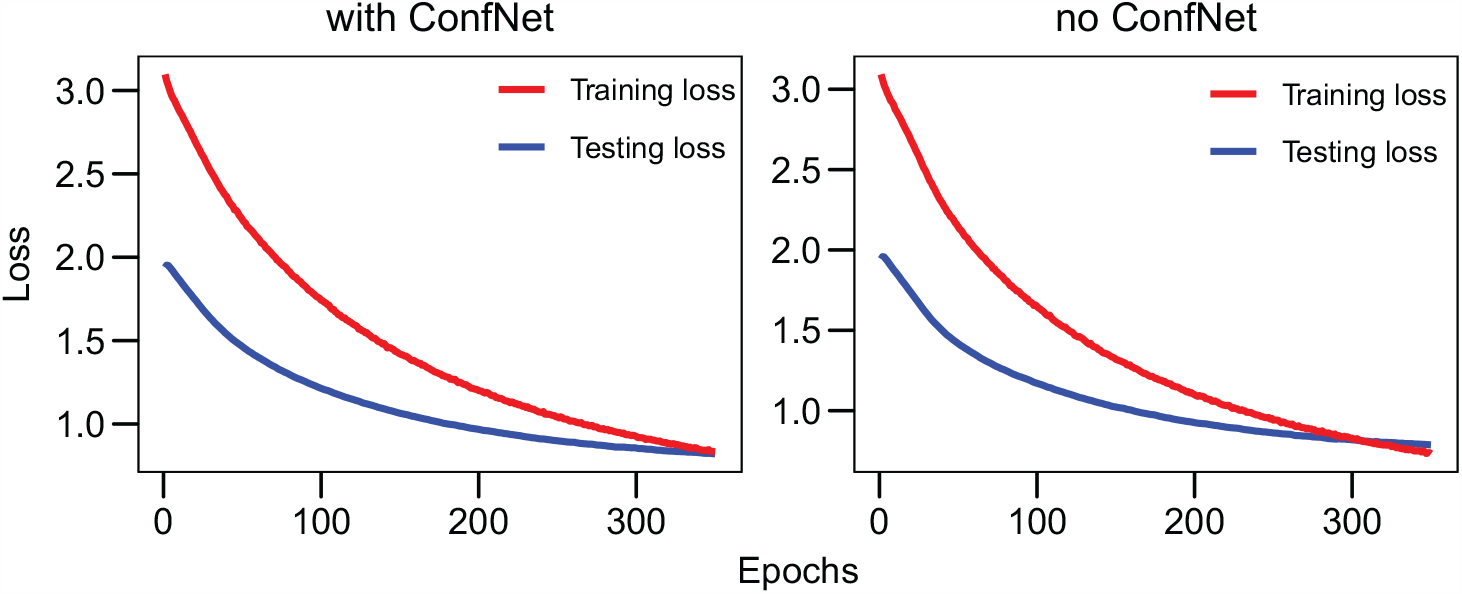
Comparison of training and testing curves with and without the ConfNet in the ROSMAP classification task.

### 3.4 Identification of Important Biomarkers

The results of our biomarker identification experiments, focusing on mRNA expression, DNA methylation, and miRNA expression, are comprehensively detailed in Table 5. This table highlights the top thirty biomarkers identified from each dataset. To validate the relevance of these biomarkers, we cross-referenced them with existing medical literature.

**Table 5.**
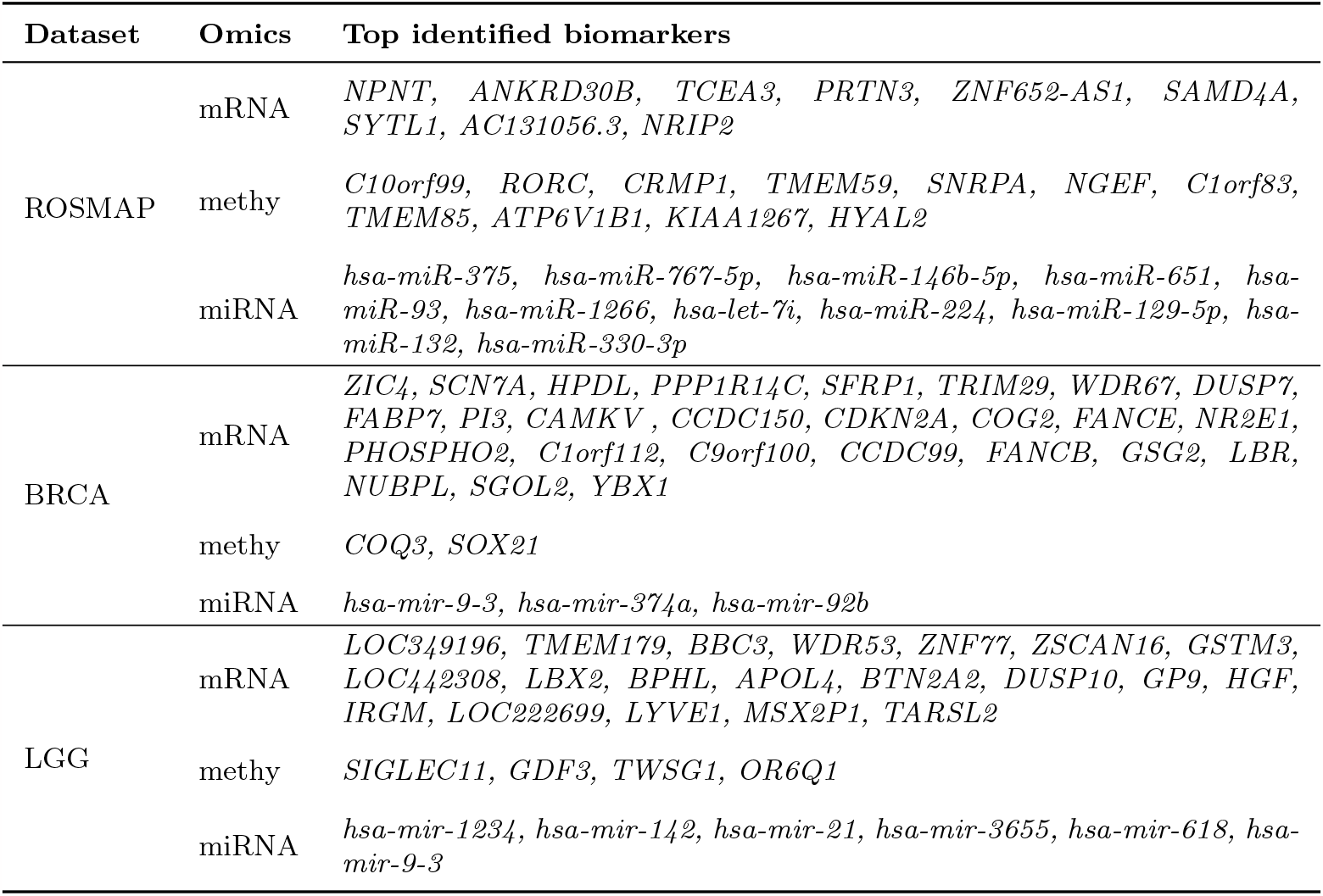
Top important biomarkers identified through our algorithm.

| Dataset | Omics | Top identified biomarkers |
| --- | --- | --- |
| ROSMAP | mRNA | *NPNT*, *ANKRD30B*, *TCEA3*, *PRTN3*, *ZNF652-AS1*, *SAMD4A*, *SYTL1*, *AC131056.3*, *NRIP2* |
|  | methy | *C10orf99*, *RORC*, *CRMP1*, *TMEM59*, *SNRPA*, *NGEF*, *C1orf83*, *TMEM85*, *ATP6V1B1*, *KIAA1267*, *HYAL2* |
|  | miRNA | *hsa-miR-375*, *hsa-miR-767-5p*, *hsa-miR-146b-5p*, *hsa-miR-651*, *hsa-miR-93*, *hsa-miR-1266*, *hsa-let-7i*, *hsa-miR-224*, *hsa-miR-129-5p*, *hsa-miR-132*, *hsa-miR-330-3p* |
| BRCA | mRNA | *ZIC4*, *SCN7A*, *HPDL*, *PPP1R14C*, *SFRP1*, *TRIM29*, *WDR67*, *DUSP7*, *FABP7*, *PI3*, *CAMKV*, *CCDC150*, *CDKN2A*, *COG2*, *FANCE*, *NR2E1*, *PHOSPHO2*, *C1orf112*, *C9orf100*, *CCDC99*, *FANCB*, *GSG2*, *LBR*, *NUBPL*, *SGOL2*, *YBX1* |
|  | methy | *COQ3*, *SOX21* |
|  | miRNA | *hsa-mir-9-3*, *hsa-mir-374a*, *hsa-mir-92b* |
| LGG | mRNA | *LOC349196*, *TMEM179*, *BBC3*, *WDR53*, *ZNF77*, *ZSCAN16*, *GSTM3*, *LOC442308*, *LBX2*, *BPHL*, *APOL4*, *BTN2A2*, *DUSP10*, *GP9*, *HGF*, *IRGM*, *LOC222699*, *LYVE1*, *MSX2P1*, *TARSL2* |
|  | methy | *SIGLEC11*, *GDF3*, *TWSG1*, *OR6Q1* |
|  | miRNA | *hsa-mir-1234*, *hsa-mir-142*, *hsa-mir-21*, *hsa-mir-3655*, *hsa-mir-618*, *hsa-mir-9-3* |

Several key findings emerged in the analysis of biomarkers within the BRCA dataset. The loss of *SFRP1* has been linked with the progression of breast cancer and a poorer prognosis in early-stage tumors [35]. Furthermore, *TRIM29* plays a role in suppressing *TWIST1* and the invasive behavior of breast cancer [36]. The expression of *C1ORF112* is notably high in both breast and cervical cancers [37]. Chen et al. [38] observed that the genetic depletion of *GSG2* marginally inhibits the growth of breast cancer cells while significantly enhancing their sensitivity to MLN8237 treatment. Additionally, the distribution of *miR-374a* in breast tumors has been examined by Li et al. [39], with implications for its role in breast cancer progression.

In the ROSMAP dataset, several biomarkers with significant implications for AD are discovered. For example, *NPNT* has been recognized as a crucial gene differentially expressed in brain tissues associated with late-onset AD [40]. Moreover, *PRTN3* has been identified as a key protective factor across various cognitive states, including dementia, mild cognitive impairment, and no cognitive impairment, and is instrumental in cognitive decline [40]. The role of *TMEM59* haploinsufficiency in reducing pathology and cognitive impairment has been well documented in the 5xFAD mouse model of AD [41]. Additionally, *Hsa-miR-375* has emerged as a novel circulating biomarker associated with extracellular vesicles in AD [42]. Furthermore, *Hsa-miR-132*, noted for its prosurvival, anti-inflammatory, and memory-enhancing functions in the nervous system, has been consistently observed to be downregulated in AD [43].

In analyzing biomarkers from the LGG dataset, several notable findings related to glioma cells have been observed. Li et al. [44] reported an increase in the expression of *GSTM3* in glioma cells compared to normal cells. Chen et al. [45] revealed that *LBX2-AS1*, a long non-coding RNA (lncRNA), is significantly upregulated in glioma, with its expression being associated with the prognosis of glioma patients. The role of *SIGLEC11* in maintaining microglia in a silent homeostatic status through sensing the intact glycocalyx of neighboring cells [46]. Zhang et al. (2020) [47] found that increased expression of *Sema3C*, which is regulated by *miR-142-5p*, indicates a poor prognosis in glioma. Additionally, Hermansen et al. (2013) [48] noted that *MiR-21* expression in the tumor cell compartment is associated with an unfavorable prognosis in gliomas.

These findings from our experiments align with existing research, thereby substantiating the robustness of our methodology in pinpointing biologically pertinent biomarkers critical for assessing disease impact.

## 4 Discussion

Our model is based on the existing shortcomings of multi-omics research, integrating auxiliary classifier-enhanced autoencoder, attention module, and the confidence network and verifying the rationality of these key components through argumentation and experimental comparison. The model not only demonstrated its state-of-the-art disease prediction ability on Alzheimer’s disease, breast cancer, gliomas, and renal cancer but also successfully detected important biomarkers for understanding disease mechanisms through feature ablation experiments.

Our contribution mainly lies in three parts. Firstly, we designed auxiliary classifiers for each omics-specific autoencoder before combining omics data. These auxiliary classifiers help train autoencoders to accurately optimize sub-representations based on task requirements, better utilize the unique features present in each omics source, and thus improve classification performance. Secondly, the attention mechanism is an effective data fusion processing method, where the model can focus more attention on omics features that significantly contribute to the classification results, further optimizing the prediction performance. Finally, the TCP criterion evaluates model confidence by comparing the predicted probabilities with real labels, thereby effectively calibrating the overconfident predictions often observed in the standard softmax output.

Our model successfully pinpoints meaningful biomarkers within each omics data type in the BRCA, ROSMAP, and LGG datasets, demonstrating robust associations with various diseases. The biomarkers identified align closely with existing medical literature findings, reinforcing the biological significance of our discoveries. The congruence of our results with established literature not only validates the efficacy of our methodology but also emphasizes the potential of these biomarkers in clinical diagnosis and their contribution to the progression of various diseases.

While our model demonstrates impressive performance, there is potential for further enhancement. First, significant opportunities remain to delve into various data fusion methodologies. For example, utilizing interaction features over basic concatenation might yield more insightful revelations regarding the interplay among features from different modalities. Another aspect worthy of exploration is the differential contributions of various model components to prediction performance. Understanding the reasons behind these varying sensitivities in different components, particularly across a range of complex diseases, presents a valuable direction for future research.

## 5 Conclusion

In this study, we have developed a multi-omics data integration framework that significantly enhances the prediction accuracy of complex diseases and demonstrates stable prediction performance across various datasets. By adeptly compressing high-dimensional data, extracting key biologically relevant features, and further leveraging omics-specific classifiers along with true class probability optimization, our framework has demonstrated superior disease classification performance compared to SOTA methods. Rigorous validation across datasets confirms the robustness and effectiveness of our model, which also serves as a potent tool for identifying critical biomarkers. These biomarkers offer profound insights into disease diagnosis and the underlying mechanisms, potentially guiding the development of targeted therapies. Thus, our work is a significant stride toward advancing precision medicine and sets the stage for subsequent research to enhance disease prediction and treatment.

## Data Availability

All data produced in the present study are available upon reasonable request to the authors
All data produced in the present work are contained in the manuscript
All data produced are available online at https://github.com/Yaolab-fantastic/MOCAT.

## Declarations

- Funding This work is supported by the National Key Research and Development Program of China (2022YFB4703500), the National Natural Science Foundation of China (62102115, 62103116), Shandong Provincial Natural Science Foundation (2022HWYQ-093), the Natural Science Foundation of Heilongjiang Province (LH2022F016), and the Fundamental Research Funds for the Central Universities (3072022TS2614).
- Conflict of interest/Competing interests The authors have no actual or potential conflicts of interest.
- Ethics approval Not applicable.
- Consent to participate Not applicable.
- Consent for publication All authors have read and agreed to the published version of the manuscript.
- Availability of data and materials All datasets used in our model training are obtained from [20] and are available at https://github.com/Yaolab-fantastic/MOCAT.
- Code availability The code is available at https://github.com/Yaolab-fantastic/MOCAT.
- Authors’ contributions Study design, Xiaohui Yao and Shan Cong; methodology, Xiaohan Jiang, Xiaohui Yao and Shan Cong; data preparation, Haoran Luo; experiments: Xiaohan Jiang and Xiaohui Yao; writing-original draft, Xiaohan Jiang, Xiaohui Yao and Shan Cong; writing-review and editing, Hong Liang, Xiufen Ye and Yanhui Wei.

## Notes

### Competing Interest Statement

The authors have declared no competing interest.

